# A Transdiagnostic Structural Brain Signature of Parkinsonian and Essential Tremor

**DOI:** 10.1101/2023.06.23.23291791

**Authors:** Christian Ineichen, Fraser Callaghan, Heide Baumann-Vogel, Fabian Büchele, Ruth O’Gorman Tuura, Christian R. Baumann, Simon J. Schreiner

## Abstract

**Background:** Parkinson’s disease (PD) and Essential Tremor (ET) are heterogeneous, yet distinct disorders. At the same time, PD and ET show overlapping features such as phenotypes with predominant tremor. These heterogeneities and overlaps pose challenges for clinical management and research and may indicate transdiagnostic, shared mechanisms for tremor.

**Objectives:** To test the hypothesis that MRI may reveal structural brain changes related to tremor phenotypes rather than diagnoses in PD and ET patients. For this, we compared regional brain volumes between three patient groups with overlapping phenotypes and distinct diagnoses: ET, PD with tremor-dominant phenotype (PD-T), and PD with non-tremor-dominant phenotype (PD-nT).

**Methods:** We studied 164 patients (18 ET, 38 PD-T, 108 PD-nT) who were evaluated for deep brain stimulation. All patients underwent structural MRI, and standardized assessment of motor symptoms. We compared regional brain volumes between groups.

**Results:** Volumes of the thalamus, pallidum, and pre-cerebellar and upper brainstem (midbrain, pons, superior cerebellar peduncle) differed across groups and were smallest in ET, intermediate in PD-T, and largest in PD-nT. Differences reached significance when comparing ET or PD-T with PD-nT but not ET with PD-T. Thalamic and brainstem volumes correlated with more severe and less levodopa-responsive tremor in PD. In contrast to the subcortical findings, cortical thickness in frontal and parietal regions was thinner in PD-nT compared to PD-T patients.

**Conclusions:** We identified tremor-related volume loss in cerebellothalamic and interconnected regions (pallidum), potentially suggesting shared mechanisms of tremor in PD and ET and pointing towards a transdiagnostic structural brain signature of tremor.

---

In movement disorders clinical practice, it is astonishing that – on the one hand - patients with distinct phenotypes can still carry the same diagnosis, as is the case in Parkinson’s disease (PD^1^), which can present without any or with predominant tremor (PD-T). On the other hand, patients with a similar, tremor-predominant phenotype, may carry distinct diagnoses, such as PD-T or Essential Tremor (ET). This heterogeneity within PD and ET and overlap between PD and ET phenotypes is in contrast to their etiological and pathophysiological concepts. Parkinson’s disease is the second most common neurodegenerative disease, with progressive nigrostriatal dopaminergic depletion and alpha-synuclein accumulation as main neuropathological features ^1^. Interestingly, phenotypic differences in PD correspond to differences in progression, non-motor burden, or even post-mortem pathology, collectively suggesting the possibility of distinct underlying etiologies ^2–4^. In contrast, ET is defined as an isolated tremor syndrome, with multiple potential etiologies and heterogeneous, mainly cerebellar, pathological features ^5, 6^. The novel concept of ET plus accounts for concomitant “soft signs” in ET patients including mild Parkinsonian signs or rest tremor that are not sufficient to diagnose a full Parkinson syndrome. Growing evidence suggests potential convergence points between PD and ET, mainly because of the higher risk and prodromal features of PD among ET patients ^7, 8^. In addition, reports of higher alpha-synuclein burden in ET have fueled the ongoing debate as to whether ET is neurodegenerative ^9, 10^ or should be considered a prodromal feature of PD ^8^, with ET plus (with Parkinsonian soft signs) representing a putative intermediate stage.

Neuroimaging studies have provided valuable insights into heterogeneity and pathophysiology: In PD, akinetic-rigid symptoms – but not tremor – closely relate to nigrostriatal dopaminergic depletion ^11^ and dysfunctional basal ganglia networks ^12^. PD tremor, on the other hand, has been linked with combined dysfunction of basal ganglia (e.g. internal pallidum) and cerebello-thalamic networks ^13, 14^. Finally, ET has been related to a selective dysfunction of the cerebellothalamic network ^13^. Furthermore, MRI studies have corroborated structural underpinnings of motor phenotypes in PD ^15–17^, such as cerebellar atrophy in tremor-dominant and cortical atrophy in akinetic-rigid subtypes. These insights suggest a pathophysiological continuum ranging from pure nigrostriatal dysfunction in akinetic-rigid PD to pure cerebellothalamic dysfunction in ET, with combined impairment in both systems in PD-T (Figure 1). In fact, recent studies showed similar connectivity changes across PD and ET, suggesting potentially shared mechanisms of tremor genesis ^18, 19^.

**Figure 1:**
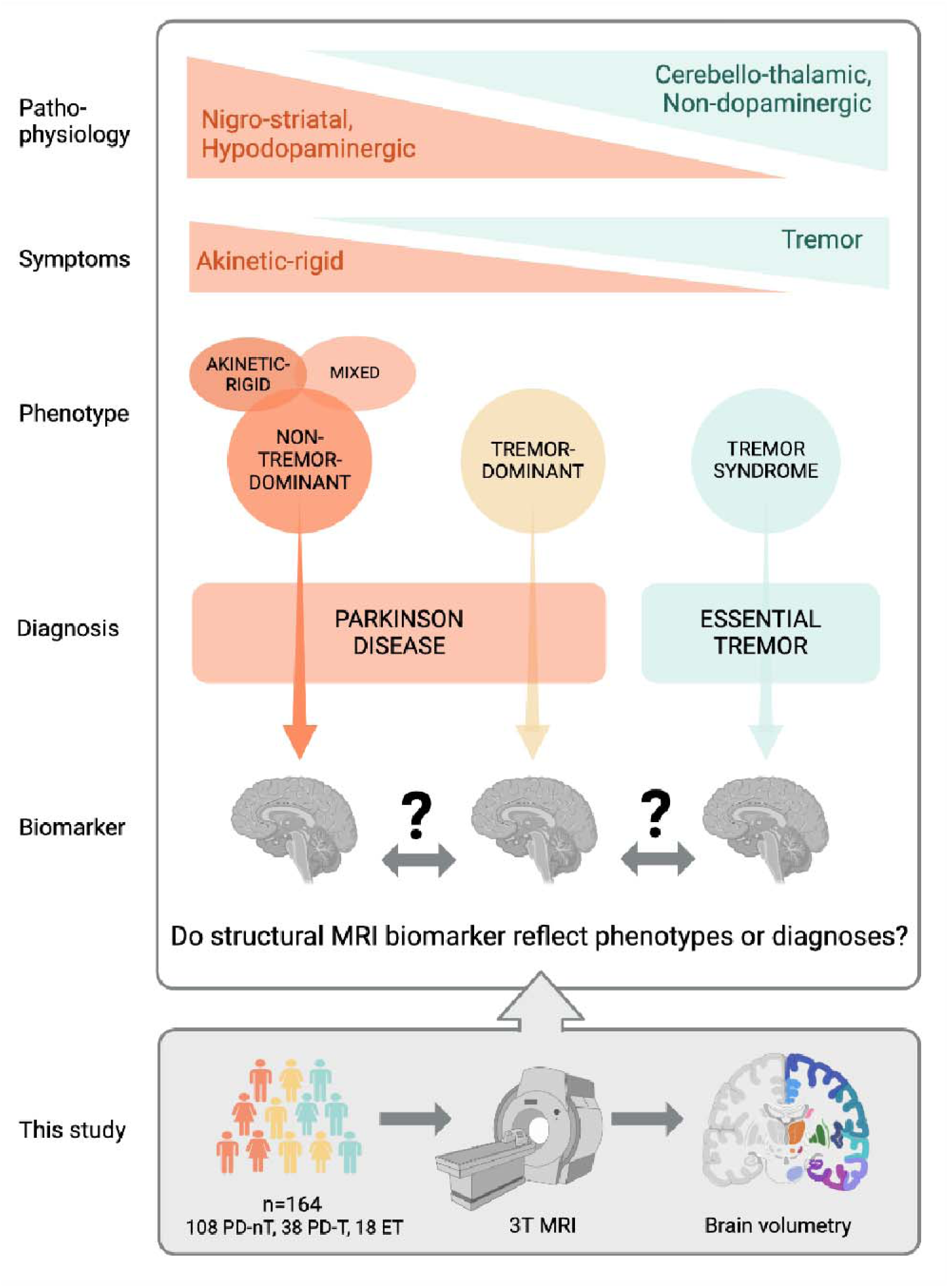
Background, study-aims and -design. Parkinson’s disease (PD) and essential tremor (ET) can hypothetically be conceptualized along a spectrum ranging from pure nigrostriatal, to combined, to pure cerebellothalamic dysfunction, resulting in distinct clinical phenotypes ranging from akinetic-rigid, to mixed, to tremor-dominant PD, and ET. Biomarkers that further characterize patients along this spectrum could improve disease understanding and treatment. Therefore, we compared structural MRI volumetrics between patients with long-standing ET or PD with (PD-T) or without tremor-dominant phenotype (PD-nT). Image created with BioRender.com.

The phenotypic within-heterogeneity and overlap between PD and ET pose major challenges for clinical management and research by interfering with diagnostic accuracy, therapeutic decisions, prognostication, and development of personalized treatments ^20, 21^. Therefore, biomarkers that characterize patients along the hypothetical PD-ET spectrum and may provide insights into underlying pathophysiology are needed.

The aim of the present study was to identify structural MRI biomarkers along a clinical spectrum from PD to ET. More specifically, we tested if regional brain volumes correspond to motor phenotypes (ET, PD with [PD-T] or without pre-dominant tremor [PD-nT]) or diagnoses (PD or ET) (Figure 1). Further, we investigated if structural brain changes correlate with clinical outcomes. Therefore, we studied clinical and regional brain volumes in patients with long-standing PD or ET, who were evaluated to undergo deep brain stimulation. We first compared regional brain volumes between ET, PD-T, and PD-nT patients. Regional volumes that differed between groups were carried further in the analysis and related to the severity and levodopa response of motor symptoms. We hypothesized that such biomarkers derived from structural MRI (1) correspond to motor phenotypes rather than diagnosis, (2) involve regions previously implicated in the pathophysiology of motor symptoms, and (3) correlate with the severity and treatment response of motor symptoms.

## Methods

### Patients

We retrospectively included consecutive patients with long-standing PD or ET who were assessed for eligibility to undergo deep brain stimulation at the Department of Neurology, University Hospital Zurich, Switzerland, between 2012 and 2018. Inclusion criteria were diagnosed PD or ET according to international diagnostic criteria ^5, 22^, available MRI with sufficient quality and documented standardized motor examinations. We collected the following clinical data: age, sex, disease duration, dopaminergic medication, Hoehn Yahr stage. In all patients, we documented the body side with more severe motor symptoms. In ET patients, upper extremity tremor was documented using the Washington Heights-Inwood Genetic Study of Essential Tremor (WHIGET) score ^23^. All PD patients underwent a levodopa challenge test, during which an experienced, specialized movement disorder nurse performed a standardized neurological examination collecting the third (motor) part of the Unified Parkinson’s Disease Rating Scale (UPDRS) during practically defined OFF (after >12 hours of drug withdrawal) and ON state (45-60 min after oral intake of levodopa). All patients gave written consent for videotaping, allowing accurate rating of motor symptoms by the examiner and an experienced movement disorder expert (C.R.B). We calculated UPDRS III total scores, and subscores for bradykinesia, rigidity, rest tremor, and postural/kinetic tremor ^24^. Based on the predominance of cardinal motor symptoms the attending neurologist assigned and documented an empirical motor subtype using the commonly used distinction of tremor-dominant, akinetic-rigid, or mixed types ^3, 4^. For the purpose of the present study, we analyzed PD phenotypes with (PD-T) or without predominant tremor (PD-nT), the latter comprising akinetic-rigid and mixed phenotypes. We retrospectively classified patients as ET or ET plus based on the presence of additional neurological soft signs [4]. For the present study, we only included patients with ET (*n* = 13) or ET plus with parkinsonian soft signs (*n* = 5). We excluded patients with ET plus with atactic (*n* = 3) or dystonic soft signs (*n* = 1), or isolated head tremor (*n* = 1). Thereby, we aimed to investigate PD and ET patients along a spectrum of akinetic-rigid and tremor symptoms (Figure 1). Out of 21 remaining ET patients, we excluded one patient because of previously implanted deep brain stimulation leads in the thalamus and one patient because of a cerebellar ischemic lesion). Additional patients had to be excluded owing to MRI quality (see section below).

### MRI data acquisition

All patients underwent MRI scanning including a T1-weighted structural MRI sequence (turbo field echo, scan mode=3D, voxel size 1×1×1 mm, TR=10.65 ms, TE=6.04 ms) using a Philips 3 Tesla Ingenia scanner (Philips Healthcare, Cleveland, OH, USA), at the Department of Radiology, University Hospital Zurich, Switzerland.

### MRI image processing

Structural T1-weighted images were analyzed using the automated image segmentation and analysis functions of FreeSurfer (version 7.1, http://surfer.nmr.mgh.harvard.edu) to perform cortical reconstruction and volumetric segmentation of the whole brain ^25^, as previously performed ^26^. In addition, we performed Bayesian segmentation of brainstem regions including the midbrain, pons, superior cerebellar peduncle (SCP), and medulla oblongata ^27^. For each participant, preprocessing included: correction of signal intensity non-uniformity in T1 images, smoothing and inflation of cortical surface data and removal of topological defects ^25^. Freesurfer outputs of interest were volume and cortical thickness. Trained analysts visually inspected segmentation outputs. Scans with insufficient quality (high noise or head motion during image acquisition; 7 PD, 1 ET) or critical lesions (1 ET with cerebellar ischemic lesion) were discarded. Cortical thickness was quality-checked by performing an outlier-analysis. No cases had to be removed.

Statistical maps of the between-group differences and the clinical associations were processed in FreeSurfer to visualize the cortical results. Data from all participants were pooled to create a study-specific group template by resampling each participant’s data into a common space, performing spatial smoothing, and then combining all scans into a single file. For the subcortical findings, a surface map was generated to display the results on an average brain. We analyzed volumes of the putamen, caudate, pallidum, thalamus, nucleus accumbens, cerebellum, and brainstem as well as its subregions medulla oblongata, pons, midbrain, and SCP. The ventral diencephalon and vessel region from the standard Freesurfer output were not included because these regions contain multiple nuclei. We averaged regional volumes across hemispheres.

### Statistical analysis

We first compared demographic and clinical data between groups of PD-T, PD-nT and ET patients using the Kruskal-Wallis-Test, with correction for multiple comparisons by the Dunn-Sydak’s post hoc test, for continuous data, or the Chi-squared test for categorical data. PD-specific data such as UPDRS III scores or levodopa equivalent dose (LED) were compared between PD-T and PD-nT patients using the Wilcoxon rank-sum test.

Subcortical brain regions: We compared subcortical volumes between groups of PD-T, PD-nT and ET patients using a multivariate analysis of covariance (MANCOVA) covarying for age, sex, and total intracranial volume (TIV). Given the number of anatomical regions considered in the analysis, reported significance values were Bonferroni-adjusted for multiple comparisons. After exclusion of 1 PD-nT patient (univariate outlier), assumptions of normality, homogeneity of variance-covariance matrices, linearity, and multicollinearity were satisfactory. In an exploratory analysis, we related subcortical brain regions with significant group differences to UPDRS III subscores of tremor and their improvement after oral intake of levodopa, using partial Spearman rank-correlation controlling for age, sex, disease duration (time from symptom onset), and TIV.

Cortical regions: Next, we tested for differences at the cortical level. We used a general linear model (GLM), to assess if cortical thickness (dependent factor) differed as a function of phenotype (ET, PD-T, PD-nT; independent factor in the design matrix), controlling for age and sex. All subjects were registered to a normalized template allowing analysis in a common domain. The spatial image distribution was smoothed with a circularly symmetric Gaussian kernel of 15 mm full width half maximum to provide normal distribution of the results. A FreeSurfer-programmed ‘Different Onset, Same Slope’ (DOSS) GLM was fitted to the spatially-normalized thickness data to create statistical maps displaying the relationships between cortical thickness and phenotypes because we wanted to examine whether a main effect of group exists independent of participant age and sex. The DOSS model constrains the slopes of any continuous variables to evolve at the same rate in all groups. Therefore, we first confirmed that there were no age-related differences on cortical thickness within the groups. Results were corrected for multiple comparisons using Freesurfer’s clusterwise correction, which considers the amount of smoothing and the number of contiguous significant points in the normalized domain. In addition, p-values were adjusted for two hemispheres using the Bonferroni correction.

## Results

Table 1 displays demographics and clinical characteristics of all subjects and each diagnostic group (PD-nT *n* = 108, PD-T *n* = 38, ET *n* = 18). ET patients were older and had a longer disease duration compared to PD-T and PD-nT patients, while age did not significantly differ between PD-T and PD-nT patients. As expected, compared to PD-nT patients, PD-T patients demonstrated more severe tremor, and less severe akinetic-rigid and axial symptoms during the OFF state. In the ON state, only tremor remained more severe in PD-T patients compared to PD-nT, presumably indicating a higher degree of levodopa-resistance of tremor compared to the other motor symptoms (Figure S1). Likewise, PD-T patients showed greater improvement of rest tremor and less improvement of axial symptoms compared to PD-nT. Total UPDRS III did not differ between groups during OFF and ON. The PD-nT group received a higher LED than the PD-T group.

**Table 1:**
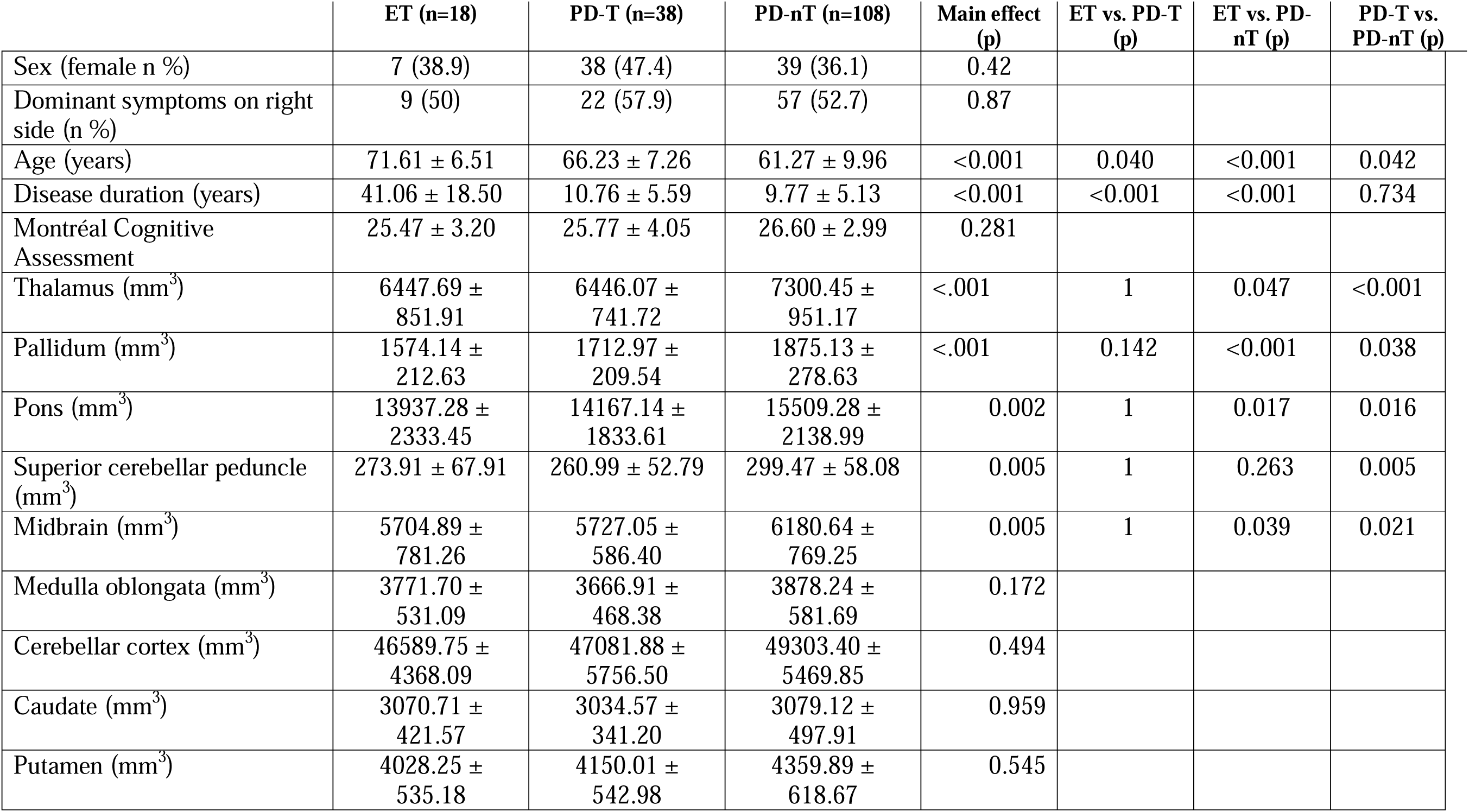
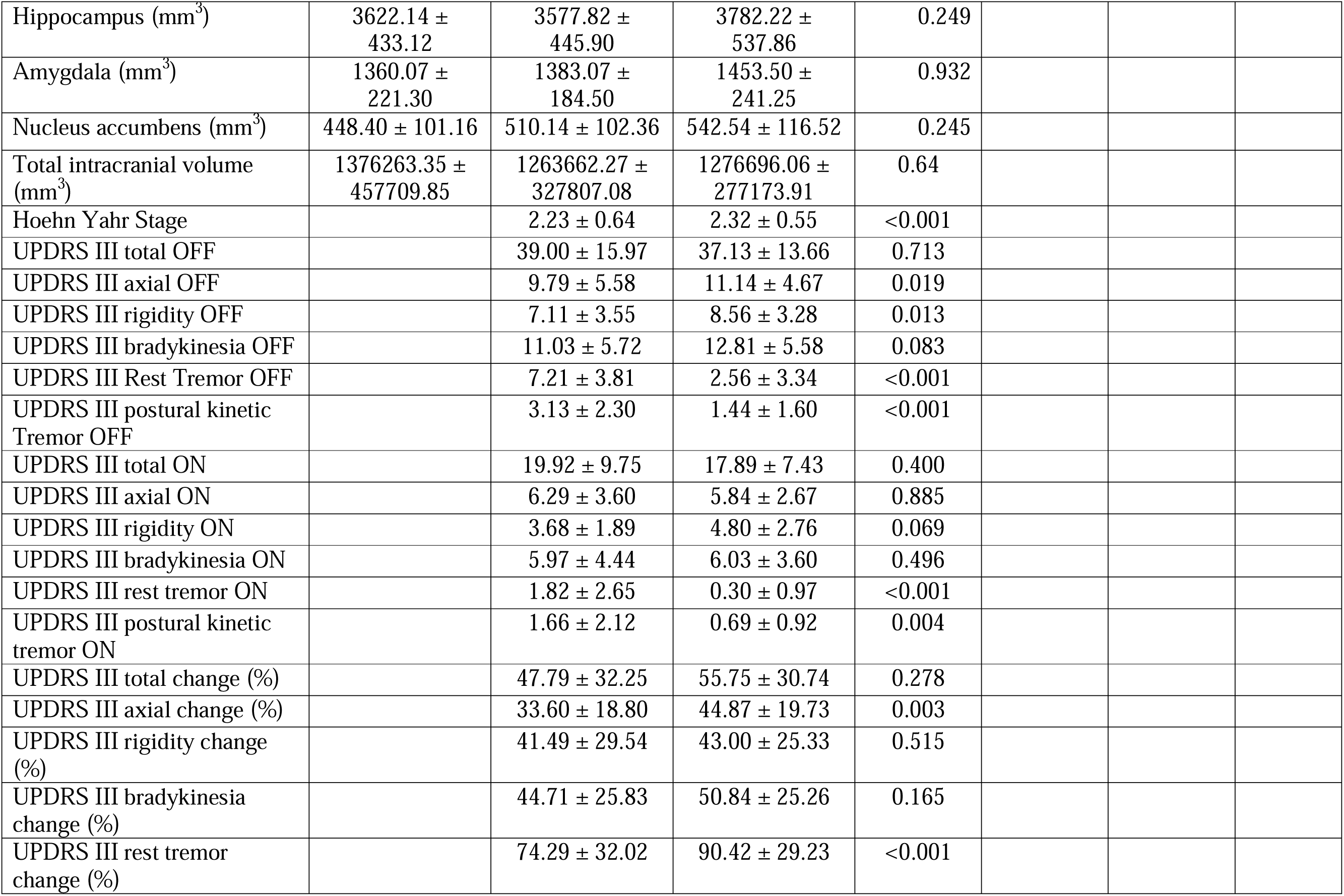
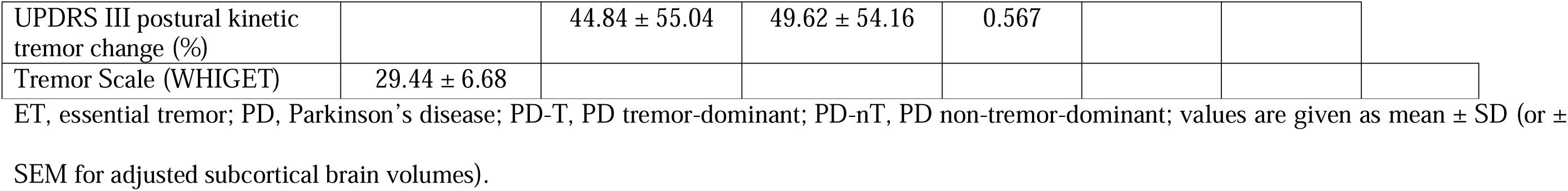
Clinical and demographic characteristics.

|  | ET (n=18) | PD-T (n=38) | PD-nT (n=108) | Main effect (p) | ET vs. PD-T (p) | ET vs. PD-nT (p) | PD-T vs. PD-nT (p) |
| --- | --- | --- | --- | --- | --- | --- | --- |
| Sex (female n %) | 7 (38.9) | 38 (47.4) | 39 (36.1) | 0.42 |  |  |  |
| Dominant symptoms on right side (n %) | 9 (50) | 22 (57.9) | 57 (52.7) | 0.87 |  |  |  |
| Age (years) | 71.61 ± 6.51 | 66.23 ± 7.26 | 61.27 ± 9.96 | <0.001 | 0.040 | <0.001 | 0.042 |
| Disease duration (years) | 41.06 ± 18.50 | 10.76 ± 5.59 | 9.77 ± 5.13 | <0.001 | <0.001 | <0.001 | 0.734 |
| Montréal Cognitive Assessment | 25.47 ± 3.20 | 25.77 ± 4.05 | 26.60 ± 2.99 | 0.281 |  |  |  |
| Thalamus (mm<sup>3</sup>) | 6447.69 ± 851.91 | 6446.07 ± 741.72 | 7300.45 ± 951.17 | <.001 | 1 | 0.047 | <0.001 |
| Pallidum (mm<sup>3</sup>) | 1574.14 ± 212.63 | 1712.97 ± 209.54 | 1875.13 ± 278.63 | <.001 | 0.142 | <0.001 | 0.038 |
| Pons (mm<sup>3</sup>) | 13937.28 ± 2333.45 | 14167.14 ± 1833.61 | 15509.28 ± 2138.99 | 0.002 | 1 | 0.017 | 0.016 |
| Superior cerebellar peduncle (mm<sup>3</sup>) | 273.91 ± 67.91 | 260.99 ± 52.79 | 299.47 ± 58.08 | 0.005 | 1 | 0.263 | 0.005 |
| Midbrain (mm<sup>3</sup>) | 5704.89 ± 781.26 | 5727.05 ± 586.40 | 6180.64 ± 769.25 | 0.005 | 1 | 0.039 | 0.021 |
| Medulla oblongata (mm<sup>3</sup>) | 3771.70 ± 531.09 | 3666.91 ± 468.38 | 3878.24 ± 581.69 | 0.172 |  |  |  |
| Cerebellar cortex (mm<sup>3</sup>) | 46589.75 ± 4368.09 | 47081.88 ± 5756.50 | 49303.40 ± 5469.85 | 0.494 |  |  |  |
| Caudate (mm<sup>3</sup>) | 3070.71 ± 421.57 | 3034.57 ± 341.20 | 3079.12 ± 497.91 | 0.959 |  |  |  |
| Putamen (mm<sup>3</sup>) | 4028.25 ± 535.18 | 4150.01 ± 542.98 | 4359.89 ± 618.67 | 0.545 |  |  |  |
| Hippocampus (mm<sup>3</sup>) | 3622.14 ± 433.12 | 3577.82 ± 445.90 | 3782.22 ± 537.86 | 0.249 |  |  |  |
| Amygdala (mm<sup>3</sup>) | 1360.07 ± 221.30 | 1383.07 ± 184.50 | 1453.50 ± 241.25 | 0.932 |  |  |  |
| Nucleus accumbens (mm<sup>3</sup>) | 448.40 ± 101.16 | 510.14 ± 102.36 | 542.54 ± 116.52 | 0.245 |  |  |  |
| Total intracranial volume (mm<sup>3</sup>) | 1376263.35 ± 457709.85 | 1263662.27 ± 327807.08 | 1276696.06 ± 277173.91 | 0.64 |  |  |  |
| Hoehn Yahr Stage |  | 2.23 ± 0.64 | 2.32 ± 0.55 | <0.001 |  |  |  |
| UPDRS III total OFF |  | 39.00 ± 15.97 | 37.13 ± 13.66 | 0.713 |  |  |  |
| UPDRS III axial OFF |  | 9.79 ± 5.58 | 11.14 ± 4.67 | 0.019 |  |  |  |
| UPDRS III rigidity OFF |  | 7.11 ± 3.55 | 8.56 ± 3.28 | 0.013 |  |  |  |
| UPDRS III bradykinesia OFF |  | 11.03 ± 5.72 | 12.81 ± 5.58 | 0.083 |  |  |  |
| UPDRS III Rest Tremor OFF |  | 7.21 ± 3.81 | 2.56 ± 3.34 | <0.001 |  |  |  |
| UPDRS III postural kinetic Tremor OFF |  | 3.13 ± 2.30 | 1.44 ± 1.60 | <0.001 |  |  |  |
| UPDRS III total ON |  | 19.92 ± 9.75 | 17.89 ± 7.43 | 0.400 |  |  |  |
| UPDRS III axial ON |  | 6.29 ± 3.60 | 5.84 ± 2.67 | 0.885 |  |  |  |
| UPDRS III rigidity ON |  | 3.68 ± 1.89 | 4.80 ± 2.76 | 0.069 |  |  |  |
| UPDRS III bradykinesia ON |  | 5.97 ± 4.44 | 6.03 ± 3.60 | 0.496 |  |  |  |
| UPDRS III rest tremor ON |  | 1.82 ± 2.65 | 0.30 ± 0.97 | <0.001 |  |  |  |
| UPDRS III postural kinetic tremor ON |  | 1.66 ± 2.12 | 0.69 ± 0.92 | 0.004 |  |  |  |
| UPDRS III total change (%) |  | 47.79 ± 32.25 | 55.75 ± 30.74 | 0.278 |  |  |  |
| UPDRS III axial change (%) |  | 33.60 ± 18.80 | 44.87 ± 19.73 | 0.003 |  |  |  |
| UPDRS III rigidity change (%) |  | 41.49 ± 29.54 | 43.00 ± 25.33 | 0.515 |  |  |  |
| UPDRS III bradykinesia change (%) |  | 44.71 ± 25.83 | 50.84 ± 25.26 | 0.165 |  |  |  |
| UPDRS III rest tremor change (%) |  | 74.29 ± 32.02 | 90.42 ± 29.23 | <0.001 |  |  |  |
| UPDRS III postural kinetic tremor change (%) |  | 44.84 ± 55.04 | 49.62 ± 54.16 | 0.567 |  |  |  |
| Tremor Scale (WHIGET) | 29.44 ± 6.68 |  |  |  |  |  |  |

ET, essential tremor; PD, Parkinson's disease; PD-T, PD tremor-dominant; PD-nT, PD non-tremor-dominant; values are given as mean ± SD (or ± SEM for adjusted subcortical brain volumes).

### Structural MRI biomarkers reflect phenotypes rather than diagnosis

Volumes of the thalamus, pallidum, and brainstem subregions differed between groups (Fig. 2 A, Table 1). Both tremor-dominant groups (PD-T and ET patients) showed smaller thalamic volumes compared to the non-tremor-dominant group (PD-nT patients) (F=9.85; p<0.001; p=0.047, respectively) (Fig. 2 B). Further, PD-T and ET patients showed smaller pallidal volumes compared to PD-nT patients (F=9.13; p=0.038; p<0.001, respectively). All subregions of the brainstem except for the medulla oblongata differed between groups (Fig. 2 B). More specifically, PD-T and ET patients had smaller volumes of the pons (F=6.34; p=0.016; p=0.017, respectively) and midbrain (F=5.54; p=0.021; p=0.039, respectively) compared to PD-nT patients. Finally, the SCP was smaller in PD-T compared to PD-nT patients (F=5.54; p=0.005).

**Figure 2:**
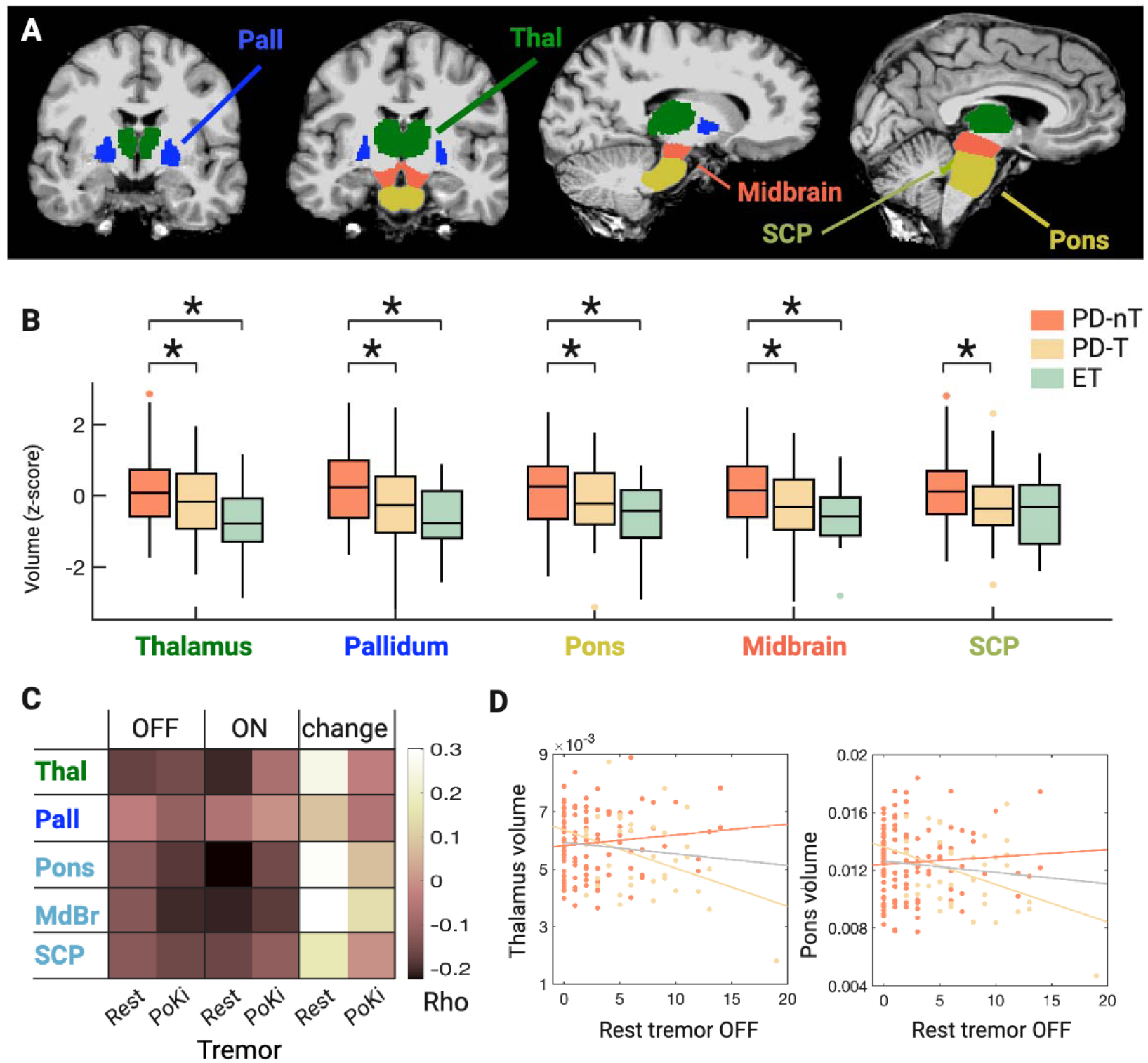
Subcortical brain signature along a phenotypic spectrum of Parkinsonian and Essential Tremor. A: Visualization of subcortical brain regions with significant group differences between patients with ET, or PD with (PD-T) or without (PD-nT) predominant tremor. B: Results revealed significant differences of group, with volumes being largest in PD-nT, intermediate in PD-T, and smallest in ET (B). C and D: Heat map and selected scatter plots of correlations of regional brain volumes with the severity of tremor during practically defined OFF and after oral intake of levodopa (ON), and the % change of tremor between OFF and ON. Volumetric data were normalized (z-transformed) in B and divided by total intracranial volume in B and D, to allow uniform visualization of brain regions of different sizes irrespective of total intracranial volume. Thal=Thalamus, Pall=Pallidum, BrSt=Brainstem, Midbr=Midbrain, SCP=Superior cerebellar peduncle, Med obl=Medulla oblongata; PKT=Postural/Kinetic Tremor. ;RT=Rest tremor; Black asterisks indicate significance at p<0.05; the gray asterisk indicates a trend at p<0.09.

Taken together, results revealed that significant differences except for one region (SCP) pertained to phenotypes rather than diagnosis, that is, tremor-predominant patients (PD-T, ET) showed smaller volumes compared to non-tremor-predominant patients (PD-nT). No significant differences emerged between PD-T and ET. Differences between ET and PD-nT were stronger than between PD-T and PD-nT, and visually volumetric differences followed a gradient, with subcortical volumes being smallest in ET, intermediate in PD-T, and largest in PD-nT (Fig. 2 B).

### Phenotype-related subcortical regions correlate with severity and drug-related improvement of tremor

Figure 2 C and D display the results of the exploratory correlation analysis relating brain volumes to tremor and its responsiveness to levodopa (see Table S1 for complete results including all rho values). Across all PD patients, smaller thalamic volume was associated with more severe and less levodopa-responsive rest tremor, with more evident associations during ON than OFF. Smaller brainstem subregions (midbrain, pons, SCP) were associated with more severe postural-kinetic tremor during OFF and ON, rest tremor during ON, and less responsiveness of rest tremor to levodopa. These associations were driven by PD-T patients and most associations were observable in PD-T but not in PD-nT patients when PD subgroups were analyzed separately. Pallidal volume showed no correlation with the severity of tremor in PD patients.

### Cortical signature of motor phenotypes across ET and PD

Using cortical thickness measures registered to a common domain, a GLM model controlling for age, sex and multiple comparisons, revealed thicker cortical thickness of two distinct regions within the superior-frontal cortex on the left hemisphere (p < 0.05, clusterwise corrected; corrected effect size, or Hedges’s g = 0.32; Fig 3, Table 2), and the precentral and superior-parietal region of the right hemisphere (p < 0.005, p < 0.01, clusterwise corrected; corrected effect sizes, or Hedges’s g = 0.13, 0.18; respectively, Fig 3, Table 2) in PD-T compared with PD-nT patients. None of the comparisons including the ET patient group revealed significant differences.

**Figure 3.**
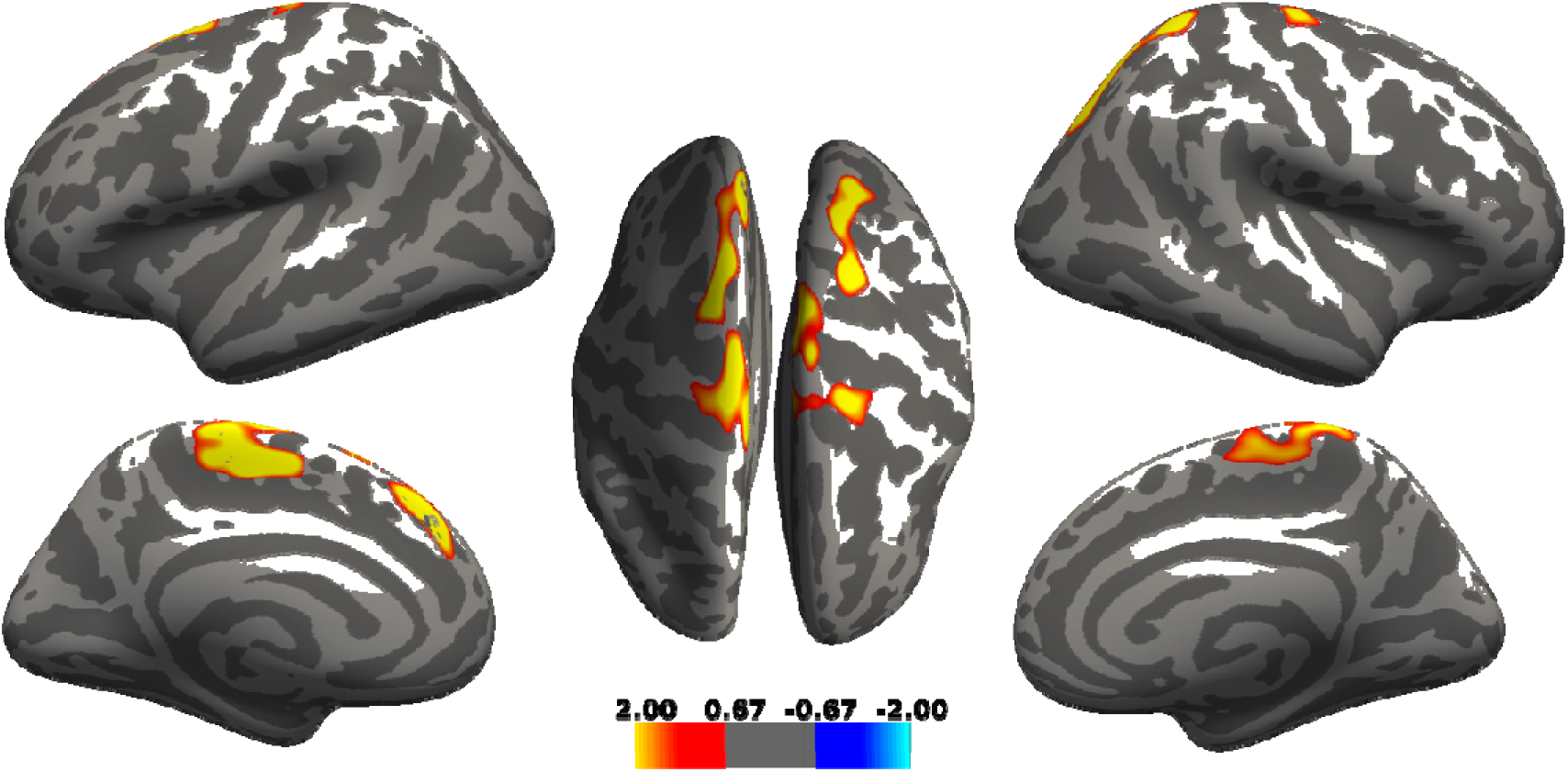
Cortical differences between patients with PD with (PD-T) or without (PD-nT) predominant tremor. Detected differences in cortical thickness between PD-T and PD-nT patients. After controlling for age and sex, thicker cortical thickness in PD-T compared to PD-nT was observed in frontal and parietal regions including two distinct subregions in the left superiorfrontal cortex, and right precentral and superiorparietal cortex (p < 0.05, clusterwise corrected). The color bar represents uncorrected significance values masked by the clusters that survived correction for multiple comparisons.

**Table 2:**
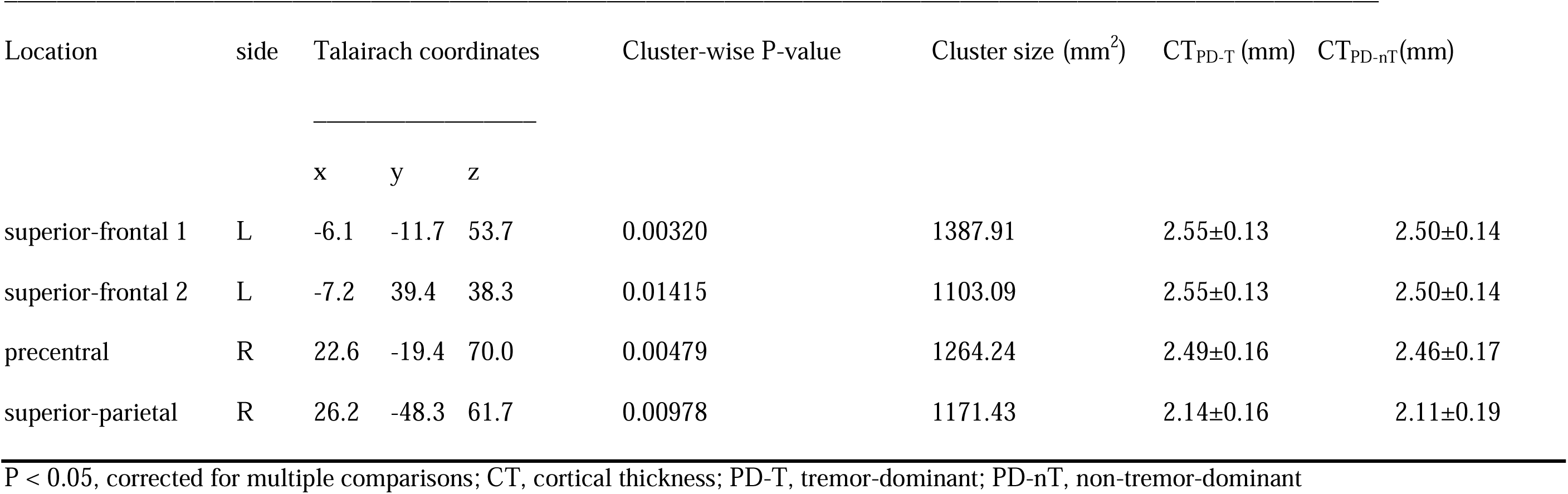
Clusters with significantly thicker cortical thickness in PD-T compared with PD-nT patients.

| Location | side | Talairach coordinates |  |  | Cluster-wise P-value | Cluster size (mm<sup>2</sup>) | CT<sub>PD-T</sub> (mm) | CT<sub>PD-nT</sub>(mm) |
| --- | --- | --- | --- | --- | --- | --- | --- | --- |
|  |  | x | y | z |  |  |  |  |
| superior-frontal 1 | L | -6.1 | -11.7 | 53.7 | 0.00320 | 1387.91 | 2.55±0.13 | 2.50±0.14 |
| superior-frontal 2 | L | -7.2 | 39.4 | 38.3 | 0.01415 | 1103.09 | 2.55±0.13 | 2.50±0.14 |
| precentral | R | 22.6 | -19.4 | 70.0 | 0.00479 | 1264.24 | 2.49±0.16 | 2.46±0.17 |
| superior-parietal | R | 26.2 | -48.3 | 61.7 | 0.00978 | 1171.43 | 2.14±0.16 | 2.11±0.19 |

P < 0.05, corrected for multiple comparisons; CT, cortical thickness; PD-T, tremor-dominant; PD-nT, non-tremor-dominant

## Discussion

We compared brain volumes across patients with ET, PD-T, or PD-nT, representing a phenotypic spectrum along which tremor decreases and akinetic-rigid symptoms increase (Figure 1). Our main finding is that volumes of the thalamus, pallidum, and pre-cerebellar and upper brainstem followed a gradient along the phenotypic spectrum, with volumes being smallest in ET, intermediate in PD-T, and largest in PD-nT. Importantly, differences reached significance when comparing tremor-dominant phenotypes (ET or PD-T) with non-tremor-dominant phenotypes (PD-nT) but not when comparing tremor-dominant phenotypes with different diagnoses (ET vs. PD-T). This finding may indicate a transdiagnostic structural signature of tremor involving the cerebellothalamic system and interconnected regions (pallidum) across PD and ET.

### Volume loss of cerebellothalamic and interconnected regions as a transdiagnostic signature of tremor

Our findings are in line with previous studies using various imaging and electrophysiological methods linking tremor to dysfunction within the cerebellothalamic system and interconnected regions in PD ^12–14^ and ET ^28–30^. Previous structural MRI studies found smaller thalamic and cerebellar volumes in ET compared to non-subtyped PD patients ^31, 32^. Our observation suggests that differences in thalamic volumes between ET and PD could be driven by PD subtype rather than PD per se. While previous studies showed tremor-related volume loss in the cerebellum ^13, 14^ and cerebellar subregions ^15, 33^, we found no group differences in cerebellar volumes. However, our study was not designed to detect localized cerebellar volume changes, because we only quantified entire cerebellar hemispheric volumes, an approach that could have limited sensitivity for more granular changes. We did, however, observe tremor-related volume loss of the pre-cerebellar and upper brainstem (SCP, pons, midbrain). Therefore, our data are indicative of tremor-related volume loss that localizes to the brainstem and thalamic portions of the cerebellothalamic system, highlighting the involvement of cerebellar pathways and regions intimately connected to the cerebellum for tremor genesis in PD and ET. Importantly, regions we identified with tremor-related volume loss did not include primary nigrostriatal projection sites such as the striatum. Thus, our observations resonate with previous studies implicating non-nigral, non-dopaminergic brainstem structures with tremor of various origins ^13^. In PD, loss of the serotonergic raphe nuclei has been linked with more severe and less levodopa-responsive tremor ^14, 34^. Moreover, quantitative susceptibility mapping MRI has indicated higher iron load in the red and dentate nucleus in PD-T compared to PD-nT patients ^35^. Interestingly, we observed that volume loss of the pre-cerebellar and upper brainstem – and to a lesser extent of the thalamus - related to both rest and postural/kinetic tremor and more so during the dopaminergic ON than OFF state. This pattern is consistent with brainstem and thalamic involvement in both dopaminergic and non-dopaminergic mechanisms of tremor ^13, 14, 34^. The absent correlation between pallidal volume and tremor in our study could be explained by the proposed indirect contribution of pallidal dysfunction to tremor, via interference with thalamocortical connections ^36^. Taken together, these data highlight the importance of non-nigral, non-dopaminergic mechanisms with particular involvement of the brainstem and cerebellothalamic system for tremor genesis in PD, similar to ET ^13, 14, 34^.

In ET, some (but not all) autopsy studies have revealed increased Lewy body inclusions in brainstem nuclei, such as the locus coeruleus ^9, 10^, suggesting that PD-like neurodegeneration could be a feature of ET at least in some patients, or that these patients could have been prone to develop PD-overlap later on ^8, 10^. Our observation of reduced brainstem volumes in ET could be interpreted as further potential evidence of a neurodegenerative component in ET, for which the clinical progression in many ET patients provides further support ^8, 10^. Given that we studied drug-resistant patients with long disease duration and evaluated for deep brain stimulation, one could speculate that neurodegeneration appears in a subgroup of ET with a more aggressive phenotype, or that long-standing severe ET phenotypes result from a so far poorly understood type of neurodegeneration involving the cerebellothalamic system. Further studies using in vivo biomarkers and postmortem histopathology are warranted to investigate biological profiles of ET subgroups and to determine if neurodegeneration discriminates severe from benign cases.

Intriguingly, the identified brain regions with tremor-related volume loss overlap with regions previously implicated with tremor-related functional changes. For example, PD-T patients showed higher metabolic activity in networks comprising the thalamus, pons, and premotor cortical regions compared to PD patients without tremor ^12^. Similarly, ET has been linked with metabolic activity in the cerebellum, brainstem, and thalamus ^28^. Recent MRI studies observed changes in structural connectivity involving the cerebellothalamic system and pallidum as a common feature of PD-T and ET compared to PD patients without tremor or healthy controls ^18, 19^. In conjunction with these former studies, our data add evidence suggesting the potential of shared mechanisms of tremor genesis across PD and ET. The anatomical agreement between regional atrophy in the present study with functional changes in previous studies allow speculating that atrophy follows functional abnormalities in cerebellothalamic pathways, yet more data is needed on this topic.

### Cortical thickness diverges from subcortical volume changes in PD

Whilst we detected smaller volumes at the subcortical level, we found a diverging pattern of thinner cortical thickness in PD-nT compared to PD-T patients, involving selected frontal and parietal areas. These cortical findings are in line with a previous study showing significantly lower gray matter volume in several cortical areas in patients with akinetic-rigid (“Postural Instability Gait Difficulty”) subtype compared to PD-T patients ^16^.

### Limitations and further implications

The study limitations pertain to the retrospective study design. Nevertheless, all data were ascertained in a structured and consecutive setting. Detailed and standardized motor assessment was carried out by the same experienced experts (C.R.B., F.B., S.J.S), but non-motor symptoms, which are important for PD phenotypes too, were not part of this study. Group sizes were imbalanced and tended to be small, especially when conducting subgroup analyses. Therefore, the study was underpowered, reflected in the relatively modest p-values calculated for cortical thickness. However, based on the narrow confidence intervals, power may have been an issue specifically for those analyses trying to isolate ET as an independent group. Further, our findings may not be generalizable to all ET patients because we studied severely affected patients and, as per study design, excluded patients outside the spectrum of tremor and akinetic-rigid symptoms (those with ET plus with soft signs of ataxia or dystonia). On the other hand, owing to the higher proportion of severe tremor our cohort may have been more suitable for studying underlying mechanisms. ET patients were older and had longer disease duration than PD patients, especially since tremor began in childhood or young adulthood in some ET patients. Therefore, it would be premature to conclude that regional volume loss is more pronounced in ET than in PD, and our findings should be interpreted with caution, although we controlled for such bias in the statistical analysis. The present study focused solely on motor phenotypes, but non-motor symptoms are essential parts of PD and ET that significantly contribute to heterogeneity and should be addressed in future studies comparing PD and ET along a spectrum.

Direct implications for clinical practice are limited at this point. Nevertheless, our data demonstrate that structural MRI can uncover meaningful biomarkers that inform about heterogeneity in PD and ET. Further, our findings confirm that structural MRI may have limited value as a diagnostic biomarker for differentiating PD-T from ET, owing to shared volumetric brain changes among tremor-dominant phenotypes irrespective of diagnosis.

### Conclusions

We found evidence of a potential transdiagnostic structural brain signature of tremor, which involves the brainstem and thalamic portions of the cerebellothalamic system and interconnected regions (pallidum) in PD and ET patients. The identified transdiagnostic structural signature of tremor is in contrast with the etiological and pathophysiological differences between PD and ET but resonates with recent studies suggesting shared mechanisms in tremor genesis as a point of convergence between PD and ET. Further studies are warranted to investigate heterogeneity and transdiagnostic mechanisms, aiming to improve diagnostic accuracy and optimal treatment choices within a personalized medicine approach in PD and ET.

## Data Availability

All data produced in the present study are available upon reasonable request to the authors.

## Acknowledgments including sources of support

The authors thank Prof. Dr. Günther Deuschl, Department of Neurology, University Hospital Schleswig-Holstein, Campus Kiel and Kiel University, Germany, and Prof. Bettina Balint, Department of Neurology, University Hospital Zurich, for their helpful comments on this article.

This research did not receive any specific grant from funding agencies in the public, commercial, or not-for-profit sectors.

## Contributors

CI: 1C, 2A, 2B, 2C, 3A

FC: 1C, 2B, 2C, 3B

HBV: 1C, 3B

FB: 3B

ROT: 2C, 3B

CB: 1A, 1B, 3B

SS: 1A, 1B, 1C, 2A, 2B, 2C, 3A

## Disclosures

### Financial Disclosures for the previous 12 months

CI: None.

FC: None.

HBV: None

FB: None.

ROT: None.

CB: Received competitive grants from the Swiss National Science Foundation, the Hochschulmedizin Zurich (Flagship Grant), the LOOP Zurich, the Novartis Foundation, and Parkinson Schweiz, and unrestricted grants from AbbVie Pharma, and Roche. Is founder and shareholder of Tosoo AG, which invests into non-pharmacological sleep modulation technologies. Received speaker honoraria from AbbVie Pharma.

SS: None.

## Ethical Compliance Statement

The study was approved by the local ethics committee (Zurich, Switzerland; BASEC 2020-02909) and patients provided written informed consent. We confirm that we have read the Journal’s position on issues involved in ethical publication and affirm that this work is consistent with those guidelines.

