## Supplementary files for "A Transdiagnostic Structural Brain Signature of Parkinsonian and Essential Tremor"

**Table S1**

| **PD all, n=146** | **RT OFF (p)** | **PKT OFF (p)** | **RT ON (p)** | **PKT ON (p)** | **RT % change (p)** | **PKT % change (p)** | **RT OFF (Rho)** | **PKT OFF (Rho)** | **RT ON (Rho)** | **PKT ON (Rho)** | **RT % change (Rho)** | **PKT % change (Rho)** |
| --- | --- | --- | --- | --- | --- | --- | --- | --- | --- | --- | --- | --- |
| Thalamus | 0.045* | 0.068 | 0.015* | 0.389 | 0.010* | 0.749 | -0.169 | -0.154 | -0.204 | -0.073 | 0.256 | -0.032 |
| Pallidum | 0.682 | 0.193 | 0.457 | 0.992 | 0.391 | 0.560 | -0.035 | -0.110 | -0.063 | -0.001 | 0.086 | -0.058 |
| Midbrain | 0.147 | 0.030* | 0.008* | 0.067 | 0.003* | 0.440 | -0.123 | -0.183 | -0.224 | -0.155 | 0.292 | 0.077 |
| Pons | 0.122 | 0.016* | 0.013* | 0.033* | 0.002* | 0.148 | -0.131 | -0.203 | -0.209 | -0.180 | 0.302 | 0.144 |
| SCP | 0.133 | 0.060 | 0.056 | 0.176 | 0.083 | 0.988 | -0.127 | -0.159 | -0.161 | -0.115 | 0.173 | -0.001 |
| **PD-T, n=38** | **RT OFF (p)** | **PKT OFF (p)** | **RT ON (p)** | **PKT ON (p)** | **RT % change (p)** | **PKT % change (p)** | **RT OFF (Rho)** | **PKT OFF (Rho)** | **RT ON (Rho)** | **PKT ON (Rho)** | **RT % change (Rho)** | **PKT % change (Rho)** |
| Thalamus | 0.073 | 0.102 | 0.169 | 0.197 | 0.305 | 0.450 | -0.316 | -0.290 | -0.245 | -0.230 | 0.184 | 0.149 |
| Pallidum | 0.834 | 0.469 | 0.070 | 0.114 | 0.104 | 0.198 | -0.038 | -0.130 | -0.319 | -0.280 | 0.288 | 0.251 |
| Midbrain | 0.082 | 0.189 | 0.049* | 0.039* | 0.078 | 0.048* | -0.307 | -0.235 | -0.346 | -0.362 | 0.311 | 0.378 |
| Pons | 0.023* | 0.111 | 0.002* | 0.015* | 0.003* | 0.036* | -0.396 | -0.282 | -0.525 | -0.420 | 0.498 | 0.399 |
| SCP | 0.428 | 0.217 | 0.318 | 0.375 | 0.539 | 0.993 | -0.143 | -0.221 | -0.179 | -0.160 | 0.111 | 0.002 |
| **PD-nT, n=108** | **RT OFF (p)** | **PKT OFF (p)** | **RT ON (p)** | **PKT ON (p)** | **RT % change (p)** | **PKT % change (p)** | **RT OFF (Rho)** | **PKT OFF (Rho)** | **RT ON (Rho)** | **PKT ON (Rho)** | **RT % change (Rho)** | **PKT % change (Rho)** |
| Thalamus | 0.976 | 0.891 | 0.914 | 0.769 | 0.331 | 0.707 | -0.003 | -0.014 | -0.011 | 0.029 | 0.124 | -0.046 |
| Pallidum | 0.526 | 0.699 | 0.053 | 0.170 | 0.123 | 0.213 | 0.063 | -0.039 | 0.191 | 0.136 | -0.196 | -0.152 |
| Midbrain | 0.893 | 0.277 | 0.342 | 0.562 | 0.160 | 0.926 | 0.013 | -0.108 | -0.095 | -0.058 | 0.179 | -0.011 |
| Pons | 0.970 | 0.161 | 0.658 | 0.352 | 0.198 | 0.624 | 0.004 | -0.139 | -0.044 | -0.093 | 0.165 | 0.060 |
| SCP | 0.847 | 0.695 | 0.183 | 0.507 | 0.116 | 0.860 | 0.019 | -0.039 | -0.132 | -0.066 | 0.200 | 0.022 |

Table shows p and partial Rho for all PD patients and subgroups; RT=rest tremor; PKT=postural-kinetic tremor; PD, Parkinson’s disease; PD-T, PD tremor-dominant; PD-nT, PD non-tremor-dominant; SCP, superior cerebellar peduncle; Rho, partial Spearman rank-correlation controlling for age, sex, and disease duration.

**Figure S1**

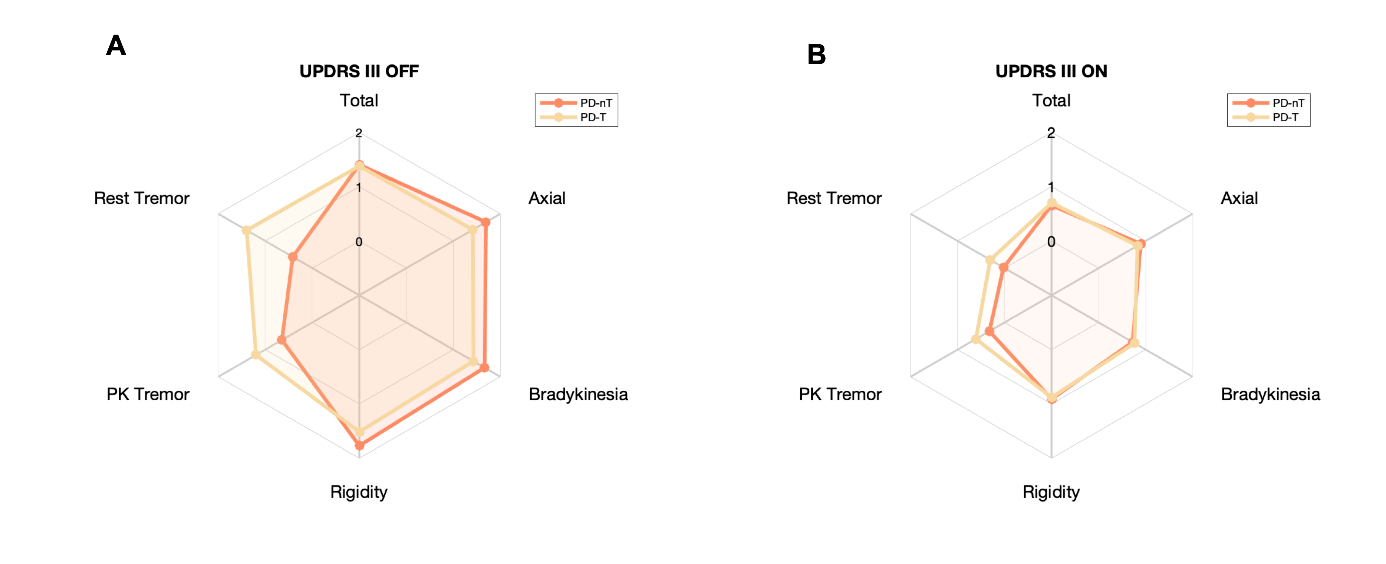

UPDRS III total and subscores during practically defined Off (OFF) and after oral intake of levodopa (ON) across PD patients with (PD-T) or without (PD-nT) predominant tremor. The UPDRS III rates the severity of each item on a scale from zero (absent) to four (severe). In this figure, we show means of total and subscores after dividing each score by the number of items contributing to that score, resulting in an average rating between zero and four. PK=Postural/Kinetic
